# Heterogeneous group of genetically determined auditory neuropathy spectrum disorders

**DOI:** 10.1101/2024.08.20.24312225

**Authors:** Anastasiia A. Buianova, Marina V. Bazanova, Vera A. Belova, Galit A. Ilyina, Alina F. Samitova, Anna O. Shmitko, Anna V. Balakina, Anna S. Pavlova, Oleg N. Suchalko, Dmitriy O. Korostin, Anton S. Machalov, Nikolai A. Daikhes, Denis V. Rebrikov

## Abstract

**Background:** Auditory Neuropathy Spectrum Disorder (ANSD) is often missed by common hearing screening tests, still accounting for up to 10% of hearing impairments. ANSD has an underlying genetic factor being mostly caused by pathogenic variants in 13 genes.

**Methods:** We examined 122 children with impaired hearing, including 102 pediatric patients (mean age was 3.7±4.1 years) with sensorineural hearing loss (SNHL) of varying severity and 20 children with a clinically confirmed ANSD (mean age was 5.65±4.63 years). For children with SNHL, we genotyped the most frequent variants (c.35delG, c.167delT, c.235delC, c.313-326del14 and c.358-360delGAG) in the *GJB2* gene using quantitative PCR. For children with a clinically confirmed ANSD, we performed the whole exome sequencing and studied the obtained variants in a custom panel of 248 genes.

**Results:** Our findings show that fifty-six (54.9%) SNHL patients carried homozygous variants in *GJB2* gene. In 12 (60%) ANSD patients, we detected variants in nucleotide sequences of the *OTOF* (25%), *CDH23, TMC1, COL11A1, PRPS1* and *HOMER2* genes (8 of which had not been previously described). Transient Evoked Otoacoustic Emissions testing revealed differences at 500 Hz (AS, p = 0.04181) and 4000 Hz (AD, p = 0.00126) between the ANSD and SNHL patient groups. The Auditory Steady-State Response (ASSR) test demonstrated significant differences at all frequencies (p < 0.01). When comparing the results obtained from the pure-tone audiometry and ASSR tests in ANSD patients revealed statistically significant differences at 500 (AD, AS) and 1000 (AD) Hz.

**Conclusions:** These findings indicate that audiologists and otorhinolaryngologists should use the ASSR test in clinical practice for differential diagnosis of auditory neuropathies. According to the survey data from the parents of hearing-impaired children, rehabilitation was more successful in SNHL patients compared to ANSD patients.

## Background

Auditory neuropathy spectrum disorder (ANSD) encompasses a wide range of hearing impairments of varying severity with rehabilitation outcomes rather difficult to predict [1]. The auditory signs of ANSD include otoacoustic emission, cochlear microphonics, and the absence of acoustic reflexes. The initial stage of audiological screening involves only the otoacoustic emissions registration [2], resulting in a delayed diagnosis of ANSD patients [3]. According to pertinent literature sources, the prevalence of ANSD varies from 1% to 10% among individuals with hearing impairment [4-6].

ANSD is often linked to prematurity, hyperbilirubinemia, congenital cytomegalovirus infection and genetic disorders [1]. However, most ANSD cases are genetically determined [7]. ANSD development is associated with variants in 13 genes [8].

Based on the localization of pathology, ANSD is classified into the pre-synaptic and post-synaptic forms [9]. The genes related to the pre-synaptic form include *OTOF, SLC17A8, CACNA1D, CABP2*. Variants in these genes disrupt inner hair cell function in the inner ear, making the affected patients potential candidates for cochlear implantation (CI). Genes linked to the post-synaptic forms of ANSD include *DIAPH3, OPA1, ATP1A3, MPZ, PMP22, NEFL, TIMM8A, AIFM1, WFS1*. Employing CI for aural rehabilitation in patients carrying the variants that cause the post-synaptic form did not prove successful [8, 10]. *OTOF* variants predominantly cause nonsyndromic ANSD [11]. Patients with OTOF-associated ANSD typically respond better to CI than to hearing aids (HA). Over 200 variants of the *OTOF* gene have been identified, with these variants accounting for congenital ANSD in more than 41% of cases in China [12].

Revealing the genetic background underlying ANSD allows for more accurate prognosis of speech development in children after CI. A review of 33 studies involving CI in children with ANSD demonstrated improvement in speech, language, and auditory parameters [13]. Predicting the efficiency of patient rehabilitation requires establishing the precise disorder etiology including an underlying genetic factor. Comparing auditory profiles among patients with different genetic variants is essential for timely diagnosis and successful auditory rehabilitation, making it a critically relevant research focus.

**The aim of our study** is to enhance the early-stage ANSD diagnosis efficacy through a comparative analysis of patient auditory profiles and genotype-phenotype matching.

## Methods

The study was conducted at the clinical base of FSBI ‘The National Medical Research Center for Otorhinolaryngology’ of the Federal Medico-Biological Agency of Russia and in the laboratories of the Russian National of Further Professional Education and laboratories of The Russian Medical Academy of Continuous Professional Education, and The National Medical Research Center For Obstetrics, Gynecology, And Perinatology Named After Academician V.I.Kulakov. All patients provided written informed consent for the sample collection, subsequent analysis, and publication thereof.

We examined 122 children with hearing impairments, including 102 pediatric patients (mean age: 3.7±4.1 years) with various degrees of sensorineural hearing loss (SNHL) and 20 children with clinically confirmed ANSD (mean age: 5.65±4.63 years).

All children underwent comprehensive clinical and audiologic assessments, including medical examinations, documentation of complaints and medical history, and auditory evaluations. Auditory assessments for SNHL patients included recording Transient Evoked Otoacoustic Emissions (TEOAEs) and Short-latency Auditory Evoked Potentials (SLAEP) using Chirp-LS signals. For ANSD patients, auditory evaluations involved recording Click signals in response to stimuli of different polarities (rarefaction and condensation phases) and Auditory Steady-State Response (ASSR) potentials. The children with ANSD were also examined using play audiometry.

After the diagnosis had been established by routine hearing tests, all patients underwent molecular genetics testing. For SNHL patients, we performed genotyping of the frequent *GJB2* variants (c.35delG, c.167delT, c.235delC, c.313-326del14 and c.358-360delGAG) using the ‘Surdogenetic’ kit (JSC DNA-Technology, Moscow, Russia) following the manufacturer’s instructions using the DT-96 thermocycler (JSC DNA-Technology, Moscow, Russia).

For children with a clinically confirmed ANSD, we carried out whole exome sequencing with subsequent analysis and data interpretation.

DNA-libraries were prepared using 500 ng of genomic DNA with the MGIEasy Universal DNA Library Prep Set (MGI Tech) following the manufacturer’s protocol. DNA fragmentation was performed via ultrasonication using Covaris S-220 resulting in the average fragment length of 250 bp. Prior to DNA fragmentation, the libraries were pooled according to the protocol described in [14] using the SureSelect Human All Exon v7 and v8 probes (Agilent Technologies), which cover the whole human exome. DNA and library concentrations were measured with Qubit Flex (Life Technologies) using the dsDNA HS Assay Kit following the manufacturer’s instructions. The quality of the prepared libraries was assessed using Bioanalyzer 2100 with the High Sensitivity DNA kit (Agilent Technologies) as per the manufacturer’s protocol. Subsequently, the libraries were circularized and sequenced in the paired-end mode using the DNBSEQ-G400 with the DNBSEQ-G400RS High-throughput Sequencing Set PE100 (MGI Tech) achieving an average coverage of 100x.

FastQ files were generated with the basecallLite software from the manufacturer (MGI Tech). The quality of the obtained sequencing data was assessed using the FastQC v0.11.9 software [15]. Based on the quality control results, the correction of raw reads was performed using the bbduk v38.96 software [16]. For each sample we conducted the bioinformatics analysis of sequencing data which included aligning reads to the human reference genome GRCh38 with bwa-mem2 v2.2.1 [17] and SAMtools v1.9 [18], identification of duplicates and obtaining the exome enrichment quality metrics using Picard v2.22.4 [19], variant calling using bcftools v1.9 [20] and Deepvariant v1.5.0 [21], variant annotation using AnnoVar [22], Intervar v2.2.2 [23] and our custom Python3 scripts for the optimization and quality improvement of the final annotation files. CNV search was performed using CNVkit v0.9.8 [24], CNV annotation was performed with ClassifyCNV v1.1.1 [25] and AnnotSV v3.2.3 [26]. After the bioinformatics analysis, we performed a final quality check with MultiQC v1.16 [27].

For this study, we assembled a panel comprising 248 genes (Table S1) associated with the diagnoses of ‘auditory neuropathy’ and ‘hearing loss’. The selection of genes was based on the Human Phenotype Ontology panels (HPO) ‘Infantile sensorineural hearing impairment (HP:0008610)’ and ‘Congenital sensorineural hearing impairment (HP:0008527)’ [28], as well as keyword searches for ‘sensorineural hearing loss’ and ‘deafness’ in the Online Mendelian Inheritance in Man (OMIM). We excluded those variants associated only with conductive hearing loss. The clinical significance of identified variants was interpreted following the ACMG criteria [29], utilizing variant databases and literature sources. The population frequencies were obtained from gnomAD v.4.0.0 [30] и RUSeq [31].

The parents of all examined children completed the survey created by the specialists of the National Medical Research Center for Otorhinolaryngology to assess the effectiveness of aural rehabilitation (Document 1).

Statistical data analysis was performed using the RStudio version 2024.03.0.

## Results

### Molecular genetic testing and the genotype-phenotype matching analysis

As shown in Fig. 1, genetic variants were observed in most patients. In our study, 56 (54.90%) SNHL patients had homozygous (47 – c.35delG, 5 – c.358-360delGAG, total 50.98%) and compound heterozygous (3 – c.35delG/c.313-326del14, 1 – c.35del/c.358-360delGAG, 3.92% in total) *GJB2* variants. Additionally, 12 (60%) ANSD patients presented with nucleotide sequence variants in various genes.

**Fig 1.**
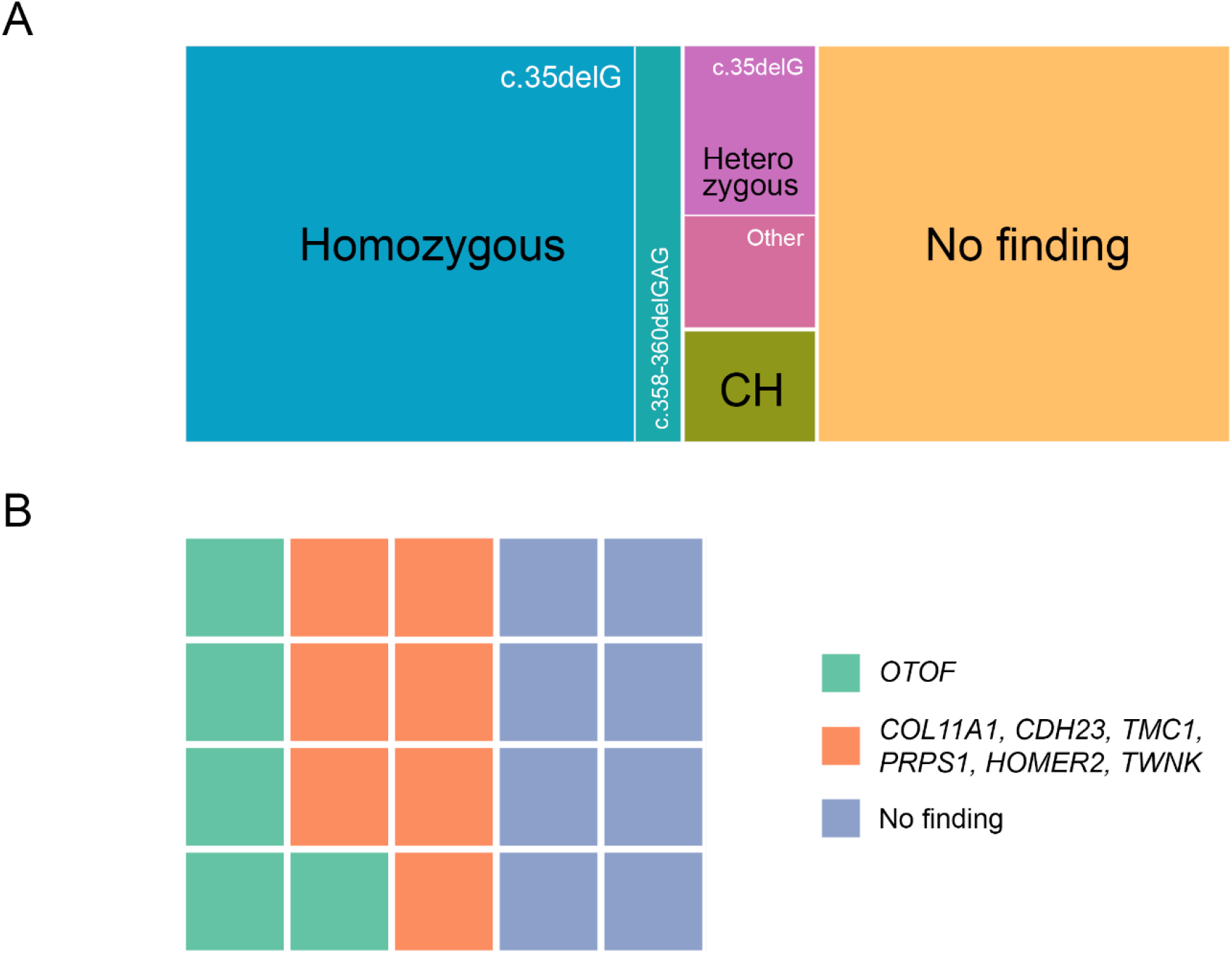
The distribution of patients depending on the presence of variants in different genes. **A** Investigated groups include conventional sensorineural hearing loss and **B** auditory neuropathy spectrum disorders. Abbreviations: CH – compound heterozygote.

Furthermore, we identified 10 carriers of *GJB2* gene variants, including 6 patients with c.35delG, and 2 each with c.358-360delGAG and c.313-326-del14 variants.

The children with ANSD and conventional SNHL were included in the main comparison group and control group, respectively. Molecular genetic testing demonstrated that the prevalence of the genetically determined hearing loss among the children undergoing treatment in the National Medical Research Center for Otorhinolaryngology was 55.74%.

Analyzing the data from the children with ANSD, we noted high levels of heterogeneity of this disorder (Table 1). Nevertheless, pathogenic variants of the *OTOF* gene were most frequently observed, occurring in homozygous or compound heterozygous states in 5 out of 20 patients.

**Table 1.**
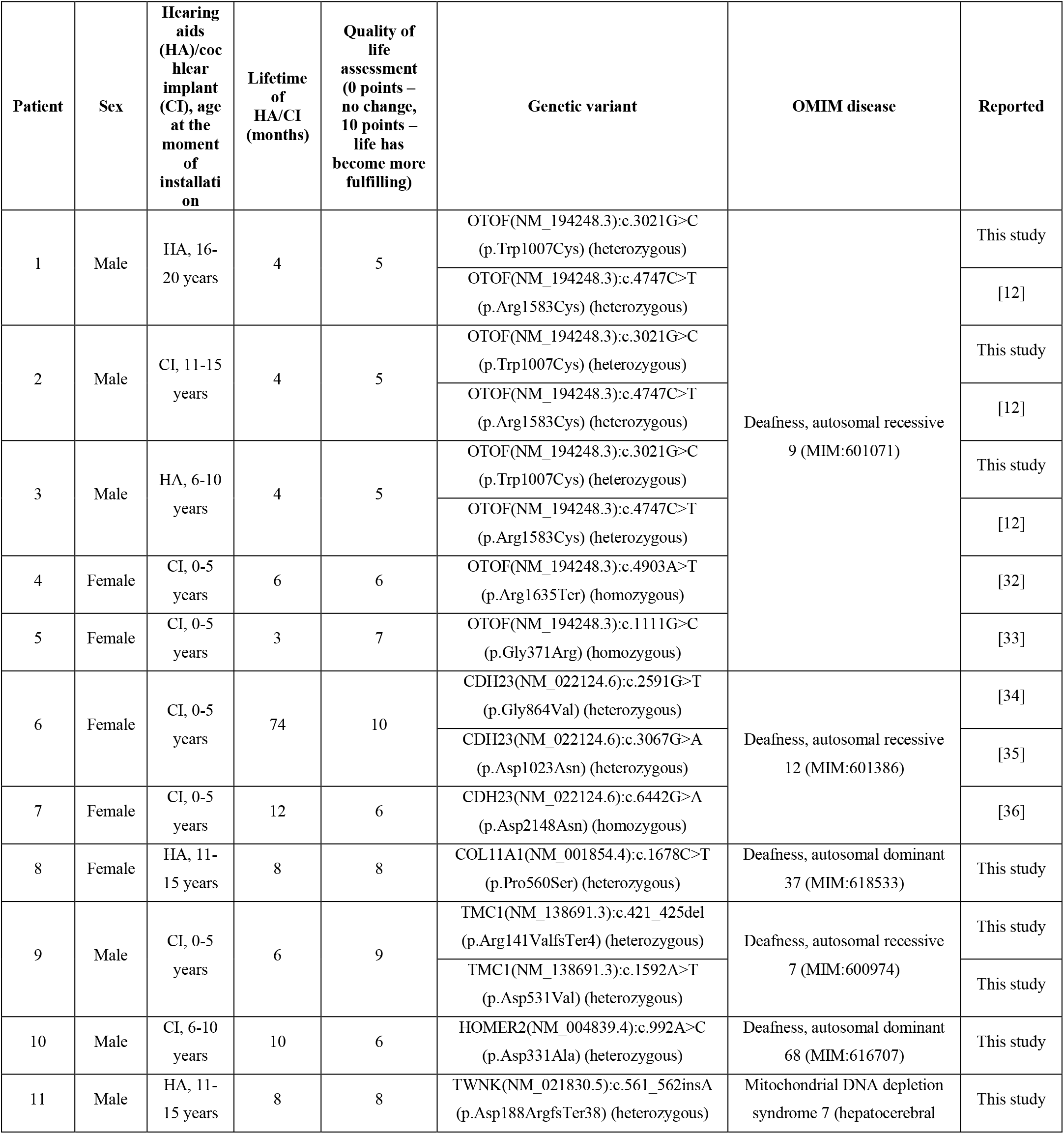
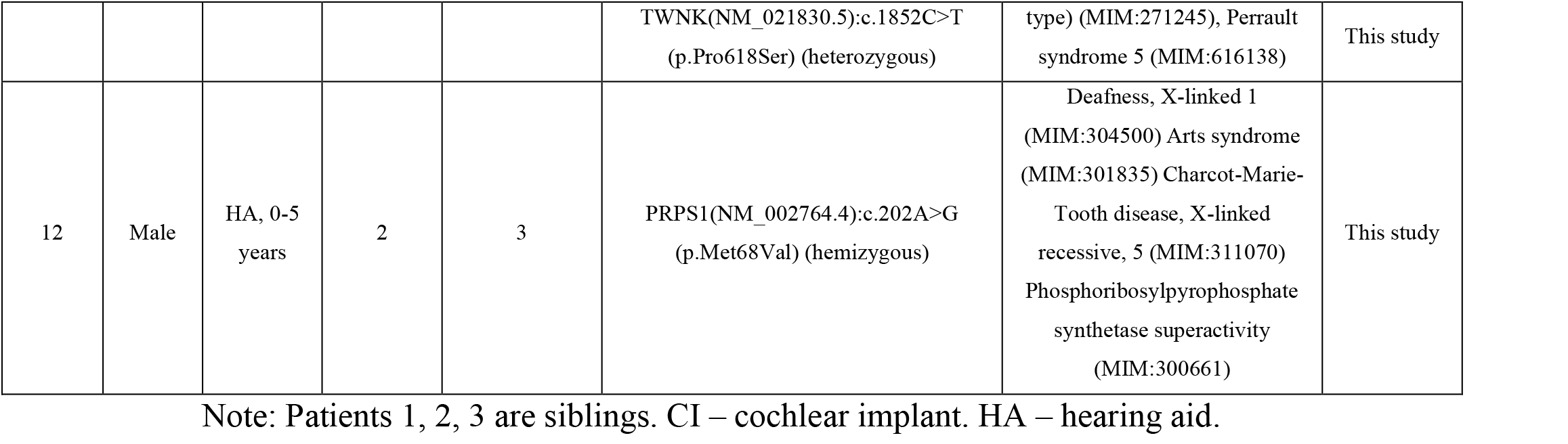
Nucleotide sequence variants in different genes among patients with auditory neuropathy spectrum disorders (ANSD) (N=12, 60% of all ANSD cases in the present study).

The extended survey results are presented in Table S2. This table includes two carriers of clinically significant variants in genes associated with ANSD: the boy with CI carried both OTOF (NM_194248.3):c.2165G>C (p.Arg722Pro) and SPNS2 (NM_001124758.3):c.310G>C (p.Ala104Pro) variants, while the girl with HA carried the CDH23 (NM_022124.6):c.5386C>A (p.Pro1796Thr) variant. All three variants have not been previously described.

Among the children with confirmed diagnoses, we identified 8 nucleotide sequence variants that have not been previously reported in the literature. We classified the following variants as likely pathogenic: OTOF(NM_194248.3):c.3021G>C (p.Trp1007Cys) (CADD score = 31), TMC1(NM_138691.3):c.1592A>T (p.Asp531Val) (CADD score = 31), TMC1(NM_138691.3):c.421_425del (p.Arg141ValfsTer4), TWNK(NM_021830.5):c.561_562insA (p.Asp188ArgfsTer38), PRPS1(NM_002764.4):c.202A>G (p.Met68Val) (CADD score = 23). Literature data reported the patient with the Charcot–Marie–Tooth disease and bilateral SNHL onset at 6-10 years of age; this patient had an altered amino acid sequence at the same position (p.Met68Leu) as identified in our patient no.12 [37]. We classified the following variants as having uncertain clinical significance: COL11A1(NM_001854.4):c.1678C>T (p.Pro560Ser) (CADD score = 28.5), HOMER2(NM_004839.4):c.992A>C (p.Asp331Ala) (CADD score = 27.4), TWNK(NM_021830.5):c.1852C>T (p.Pro618Ser) (CADD score = 22.5). Despite the p.Pro618Ser variant being predicted as benign, it is located in the functional SF4 helicase domain, which contains 97 missense/in-frame variants, including 36 pathogenic variants, 58 variants of unknown significance, and only 3 benign variants.

Pathogenic variants in the *OTOF* gene were transmitted from mothers to patients 4 and 5 (fathers were not available for genetic analysis). Patient 9 inherited the *TMC1* p.Arg141ValfsTer4 and p.Asp531Val variants from his mother and father, respectively. Patient 12 inherited the PRPS1 variant p.Met68Val from his mother.

Patient 5 also harbored a previously undescribed variant TECTA(NM_005422.4):c.4966A>G (p.Met1656Val) in a heterozygous state, associated with deafness, autosomal dominant 8/12 (MIM:601543), and deafness, autosomal recessive 21 (MIM:603629). However, according to CADD, this variant was classified as benign (score = 22.6).

Patient 11 was monitored throughout infancy for breath-holding spells and perinatal brain injury. The patient was diagnosed with delayed speech development. Five years after the onset of the condition, the hearing loss was observed, initially unilateral and later bilateral. The patient is under the care of an otorhinolaryngologist and audiologist with a diagnosis of stage 2 bilateral mixed (predominantly sensorineural) hearing loss. Additionally, mild myotonic syndrome has been diagnosed by a neurologist. Stage 2 bilateral SNHL is present, along with speech impairment, asthenic syndrome, and behavioral issues. A psychiatrist diagnosed the patient with F06.7 (ICD-10).

### Hearing Test Results

In our study, a comparative analysis of the signal-to-noise ratio during the registration of TEOAEs (Fig. 2A, Table 2) revealed a statistically significant difference at 500 Hz (AS) and 4000 Hz (AD) between the ANSD patients and control children. The least pronounced difference was observed at 500 Hz (AD), since both groups showed negative signal-to-noise ratios.

**Table 2.**
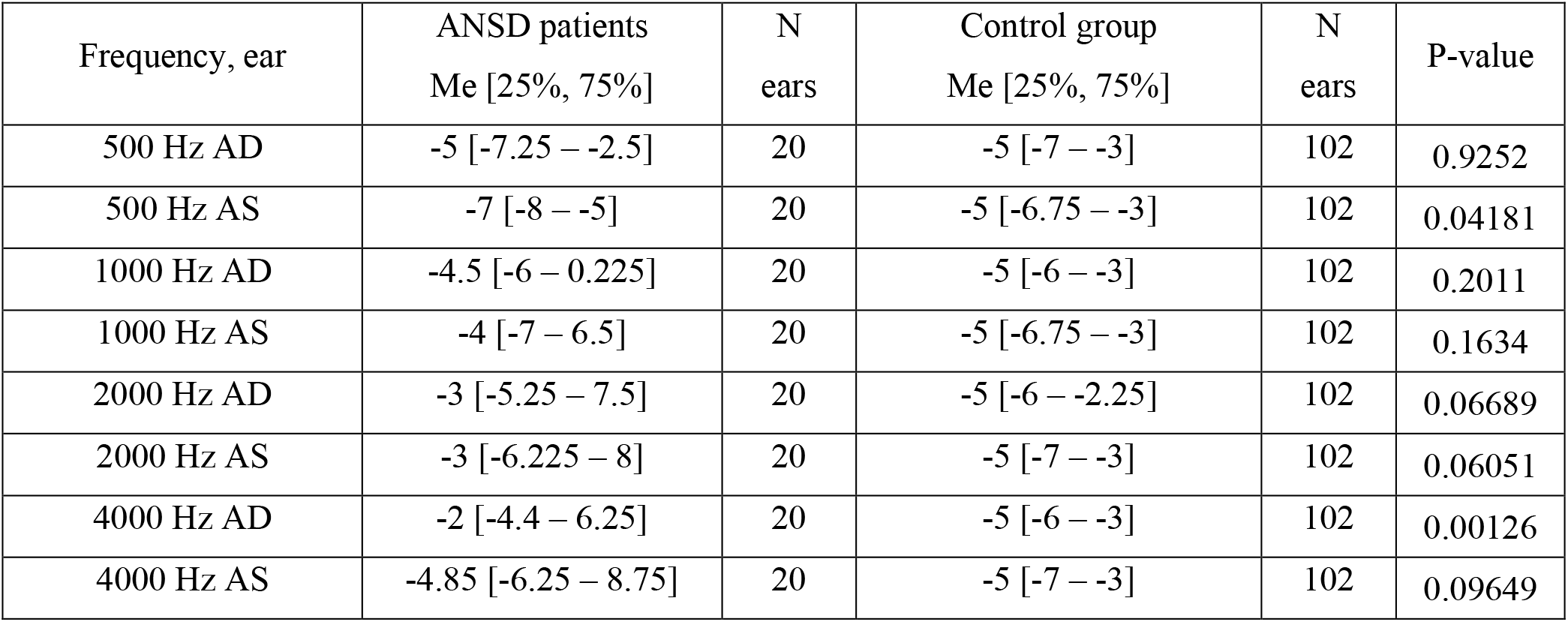
Comparative characterization of signal-to-noise ratios during the registration of Transient Evoked Otoacoustic Emissions (TEOAEs) in patients with auditory neuropathy spectrum disorders (ANSD) and control group patients. Mann-Whitney U test.

**Fig 2.**
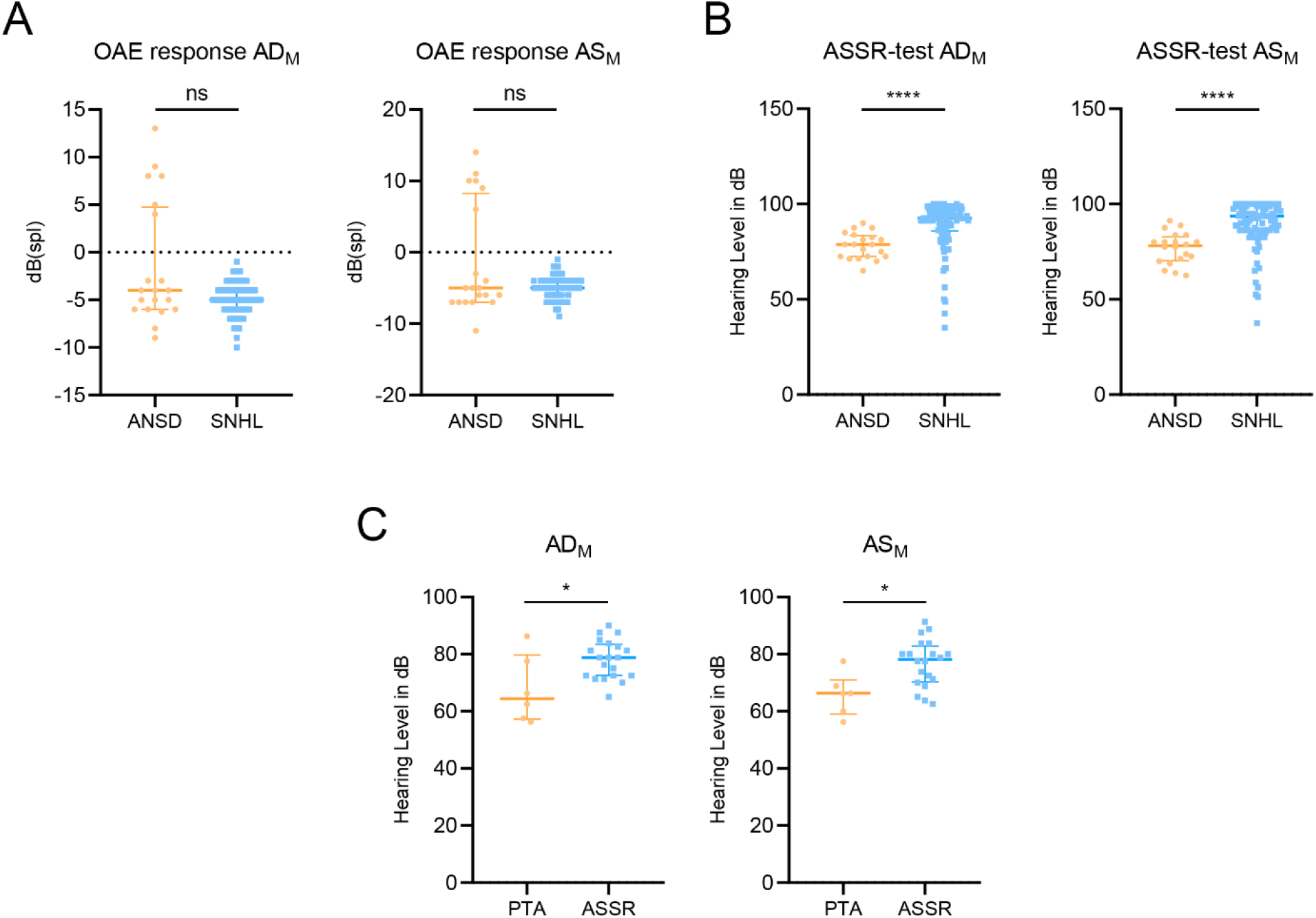
Comprehensive audiologic assessments. **A** Comparative analysis of mean signal-to-noise ratios during transient-evoked otoacoustic emissions (TEOAEs) registration in patients from the main and control groups. **B** Comparative analysis of mean Auditory Steady-State Response (ASSR) test values in patients from the main and control groups. **C** Comparison of mean results from pure-tone audiometry (PTA) and ASSR tests in patients with auditory neuropathy spectrum disorders (ANSD). Abbreviations: SNHL – sensorineural hearing loss; OAE – otoacoustic emissions. Mann-Whitney U test. p ≤ 0.05 is designated as ‘*’, p < 0.0001 – as ‘****’; ns – not significant (p > 0.5).

Table 3 presents the comparative analysis of the ASSR test results in patients from the main and control groups, revealing significant differences at all frequencies, particularly notable at 2000 Hz and 4000 Hz (AD and AS). As shown in Fig. 2B, ANSD patients exhibited lower and more varied detection threshold values compared to the control group.

**Table 3.** A comparison of the Auditory Steady-State Response (ASSR) test results between auditory neuropathy spectrum disorders (ANSD) and control patients. Mann-Whitney U test.

| Frequency, ear | ANSD patients<br>Me [25%, 75%] | N<br>ears | Control group<br>Me [25%, 75%] | N<br>ears | P-value |
| --- | --- | --- | --- | --- | --- |
| 500 Hz AD | 75 [65 – 80] | 20 | 90 [80 – 95] | 102 | $4.079 \times 10^{-5}$ |
| 500 Hz AS | 80 [75 – 85] | 20 | 90 [85 – 100] | 102 | 0.009459 |
| 1000 Hz AD | 80 [75 – 90] | 20 | 95 [85 – 100] | 102 | 0.001726 |
| 1000 Hz AS | 80 [73.75 – 82.5] | 20 | 95 [90 – 100] | 102 | $1.091 \times 10^{-5}$ |
| 2000 Hz AD | 80 [78.75 – 90] | 20 | 95 [86.25 – 100] | 102 | $1.465 \times 10^{-6}$ |
| 2000 Hz AS | 77 [70 – 80] | 20 | 90 [90 – 100] | 102 | $1.108 \times 10^{-6}$ |
| 4000 Hz AD | 72.5 [68.75 – 90] | 20 | 90 [85 – 100] | 102 | $1.935 \times 10^{-5}$ |
| 4000 Hz AS | 70 [60 – 85] | 20 | 95 [85 – 100] | 102 | $2.989 \times 10^{-7}$ |

Comparing the right and left ears within the ANSD patient group, the registration of peak I of click-evoked auditory brainstem responses (ABR) and auditory thresholds measured by the ASSR test showed symmetrical results, with statistically insignificant differences.

In ANSD patients, results from pure-tone audiometry (PTA) indicated lower thresholds compared to those recorded during the ASSR test (Fig. 2C). Comparative analysis of metrics obtained from the ASSR test and PTA revealed statistically significant differences at frequencies of 500 Hz (AD and AS) and 1000 Hz (AD) within the ANSD patient group (Table 4).

**Table 4.** Pure-tone audiometry (PTA) and the Auditory Steady-State Response (ASSR) test comparison of the right and left ear in auditory neuropathy spectrum disorders patients. Mann-Whitney U test.

| Frequency, ear | PTA test<br>Me [25%, 75%] | N<br>ears | ASSR test<br>Me [25%, 75%] | N<br>ears | P-value |
| --- | --- | --- | --- | --- | --- |
| 500 Hz AD | 57.5 [47.5 – 60] | 6 | 75 [65 – 80] | 20 | 0.003783 |
| 500 Hz AS | 50 [46.25 – 53.75] | 6 | 80 [75 – 85] | 20 | 0.00083 |
| 1000 Hz AD | 65 [61.25 – 76.25] | 6 | 80 [75 – 90] | 20 | 0.0383 |
| 1000 Hz AS | 67.5 [57.5 – 70] | 6 | 80 [73.75 – 82.5] | 20 | 0.05242 |
| 2000 Hz AD | 72.5 [66.25 – 82.5] | 6 | 80 [78.75 – 90] | 20 | 0.1437 |
| 2000 Hz AS | 70 [61.25 – 75] | 6 | 77.5 [70 – 80] | 20 | 0.1946 |
| 4000 Hz AD | 72.5 [70 – 82.5] | 6 | 72.5 [70 – 90] | 20 | 0.9754 |
| 4000 Hz AS | 80 [76.25 – 80] | 6 | 70 [60 – 85] | 20 | 0.4065 |

### Quality of life assessment

Patients were stratified based on the use of rehabilitation tools: 11 ANSD patients and 83 SNHL patients used CI, while 9 ANSD patients and 19 SNHL patients used hearing aids. In both groups, patients predominantly used CI.

According to the survey (Document 1), SNHL patients had been using technical devices longer (20.37±16.4 months) than ANSD patients (14.35±13.1 months).

Quality of life assessment with a 10-point rating scale showed that the rehabilitation of SNHL patients proved to be more successful as compared to ANSD patients.

## Discussion

In our study, the genetically determined hearing loss accounted for 74% of all children undergoing both treatment and monitoring at the FSBI ‘The National Medical Research Center for Otorhinolaryngology’ of the Federal Medico-Biological Agency of Russia, with ANSD patients comprising 17.65% of these cases. The most common genetic cause of ANSD observed was variants in the *OTOF* gene, consistent with the existing literature, occurring in 25% of cases in our study.

From our findings, only a subset of the identified genes (*OTOF, TWNK, PRPS1*) could be directly linked to ANSD. For instance, one patient with compound heterozygosity for the p.Arg1583His and p.Gln1883* variants in the *OTOF* gene underwent cochlear implantation at the age of 20 months. After five years of monitoring, their average pure tone thresholds at 500, 1000, 2000, and 4000 Hz increased by 25–37.5 dB [38]. Another patient with the Perrault syndrome, associated with *TWNK* variants and onset of hearing loss at 5.5 years, received a CI at 6.5 years old. After 1 year, the average hearing threshold increased from 98.75 dB to 38.25 dB; the vowel, consonant, disyllable, and tone recognition scores in the quiet field were 36, 36, 36, and 56%, respectively [39]. In this patient and another patient carrying compound heterozygous variants of the *TWNK* gene, CI improved CAP and SIR scores.

The contribution of the *COL11A1, CDH23, TMC1, HOMER2* genes to ANSD development has not been previously described, which emphasizes that we still lack profound knowledge of this disease. CI was found to be completely ineffective for a German patient with the heterozygous variant COL11A1(NM_080629.2):c.2644C>T (p.Arg882Trp) [40]. Conversely, patients with *CDH23* variants demonstrated improvements in hearing and speech outcomes post-CI [41], including those with the CDH23(NM_022124.6):c.2591G>T (p.Gly864Val) variant in a compound heterozygous state, similar to findings in patient 6 [34]. Although the CDH23(NM_022124.6):c.6442G>A (p.Asp2148Asn) variant is prevalent in Europe with 138 healthy heterozygous individuals reported [30], no cases of CI in patients with this variant have been documented. Excellent clinical outcomes were observed with CI in two half-siblings with compound heterozygosity at the TMC1 gene (p.Arg34 and p.Trp321*) and in a patient with variants p.Arg389* and p.Arg512* [42]. *HOMER2*-associated deafness is extremely rare, with the only described variants being p.Arg196Pro, p.Met281Hisfs*9, p.Pro278Alafs*10. The patients carrying these variants did not undergo CI [43]. As whole genome and whole exome sequencing become more accessible, the development of multigene panels, including ‘hearing loss’ panels with dozens of genes associated with hearing loss, presents a promising approach for detecting rare forms of congenital hearing loss in ANSD patients.

TEOAEs revealed a statistically significant difference between ANSD and SNHL patients at 500 (AS) and 4000 Hz (AD). Notably, patients with *OTOF* gene variants continued to exhibit TEOAEs responses regardless of age, consistent with the existing literature [44]. In contrast, TEOAEs were absent in other patients, and the signal-to-noise ratio was negative, leading to no significant differences between the groups.

The ASSR test demonstrated a statistically significant difference between ANSD and SNHL patient groups, with ANSD patients showing lower parameter values. Given the clinical and auditory characteristics of ANSD, we conducted a comparative analysis of PTA and ASSR test results, revealing differences only at 500 Hz (AD, AS) and 1000 Hz (AD). However, in our study, some patients couldn’t be examined by PTA because of their age or inability to understand the task.

Therefore, our analysis underscores the importance of employing both ASSR tests and PTA for differential diagnosis of ANSD and SNHL in clinical practice by audiologists-otorhinolaryngologists.

According to a survey of the parents of the children with hearing impairment, rehabilitation outcomes were more successful in SNHL patients compared to ANSD patients. Currently, we cannot evaluate the outcomes of aural rehabilitation of ANSD patients, since not all patients have been using HA/CI for at least 5 years. We hope to report new meaningful results in our further research.

## Supporting information

Document 1 (docx). The screening survey (younger than 18 years).

## Data Availability

The sequence data are generated from patient samples and therefore are available under restricted access.

## Abbreviations

ABR: Auditory Brainstem Responses
AD: Right ear
ANSD: Auditory Neuropathy Spectrum Disorder
AS: Left ear
ASSR: Auditory Steady-State Response
CH: Compound heterozygote
CI: Cochlear implant
HA: Hearing aid
HPO: Human Phenotype Ontology
OMIM: Online Mendelian Inheritance in Man
PTA: Pure-tone audiometry
SLAEP: Short-latency Auditory Evoked Potentials
SNHL: Sensorineural hearing loss
TEOAEs: Transient Evoked Otoacoustic Emissions

## Supporting Information

Additional file 1:

Table S1 (xlsx). The gene panel ‘Deafness’.

Table S2 (xlsx). Results of the survey of the patients with genetically confirmed ANSD and the carriers of the ANSD genes variants.

Additional file 2:

Document 1 (docx). The screening survey (younger than 18 years).

## Ethics approval and consent to participate

This project was approved by local ethical committee of the FSBI ‘The National Medical Research Center for Otorhinolaryngology of the Federal Medico-Biological Agency of Russia’ on the 26th of April 2021 (Protocol No. 02/21), and all participants provided written informed consent prior to data collection.

## Consent for publication

Not applicable.

## Author Contributions

NAD, ASM, ASM and DVR conceived and designed the study. MVB, AVB, ASM and MVB recruited the patients. AAB developed the genetic panel for analyzing patient annotation files, carried out the clinical interpretation of these data, conducted statistical analyses and created the graphs. VAB, AOS, AFS and GAI isolated DNA from the patients’ blood and performed exome sequencing. ASP and ONS developed the bioinformatics pipeline. NAD and AVB provided critical feedback on the study. NAD, ASM, DOK and DVR supervised the study. All authors read and approved the final manuscript.

## Acknowledgements

Not applicable.

