## Supplementary material for "Heterogeneous group of genetically determined auditory neuropathy spectrum disorders": Document 1 (docx). The screening survey (younger than 18 years).

**Screening Diagnostic Questionnaire (for children)**

| ***Full Name ___________________________________________________*** | | | | | |
| --- | --- | --- | --- | --- | --- |
| ***Age (in full years) ___________________________________*** | | | | | |
| ***Duration of use of the rehabilitation device: ______________________*** | | | | | |
| **Educational segment (underline the correct one):** not organized (home), general daycare, general school, special school for hearing-impaired, university, vocational institution. | | | | | |
| ***Mark the correct answer.*** | | **Yes (0 points)** | **No (2 points)** | | **Sometimes (1 point)** |
| **1.** | Does your child have difficulties in communication (with CI/HA)? |  |  | |  |
| **2.** | Does your child feel uncomfortable wearing a rehabilitation device? |  |  | |  |
| **3.** | Does your child experience headaches when wearing CI or HA for a long time? |  |  | |  |
| **4.** | Does your child have difficulties talking on the phone with CI/HA (if using a phone)? |  |  | |  |
| ***Mark the correct answer in the corresponding cell.*** | | **Yes (2 points)** | | **No (0 points)** | |
| **5.** | Does your child undergo rehabilitation courses? |  | |  | |
| **6.** | Does your child study with a speech therapists/deaf teacher? |  | |  | |
| ***Please answer the following questions.***  **7.** What time per a day does your child explore CI or HA? ___________________________________________  **8.** How often does your child visit an audiologist? ___________________________________________ | | | | | |
| **9. *(Rate on a 10-point scale where 0 – no change and 10 – life has become brighter and more fulfilling)***  Your child's life has changed since they started wearing HA or CI.  0 1 2 3 4 5 6 7 8 9 10 | | | | | |
